# Long-read sequencing identifies copy-specific markers of *SMN* gene conversion in spinal muscular atrophy

**DOI:** 10.1101/2024.07.16.24310417

**Authors:** M.M. Zwartkruis, M.G. Elferink, D. Gommers, I. Signoria, L. Blasco-Pérez, M. Costa-Roger, J. van der Sel, I.J. Renkens, J.W. Green, J.V. Kortooms, C. Vermeulen, R. Straver, H.W.M. van Deutekom, J.H. Veldink, F. Asselman, E.F. Tizzano, R.I. Wadman, W.L. van der Pol, G.W. van Haaften, E.J.N. Groen

## Abstract

The complex 2 Mb *survival motor neuron (SMN)* locus on chromosome 5q13, including the spinal muscular atrophy (SMA)-causing gene *SMN1* and modifier *SMN2*, remains incompletely resolved due to numerous segmental duplications. Variation in *SMN2* copy number, presumably influenced by *SMN1* to *SMN2* gene conversion, affects disease severity, though *SMN2* copy number alone has insufficient prognostic value due to limited genotype-phenotype correlations. With advancements in newborn screening and *SMN*-targeted therapies, identifying genetic markers to predict disease progression and treatment response is crucial. Progress has thus far been limited by methodological constraints. To address this, we used targeted nanopore long-read sequencing to analyze copy-specific variation in *SMN* and neighboring genes. In 25 healthy controls, we identified single nucleotide variants (SNVs) specific to *SMN1* and *SMN2* haplotypes that could serve as gene conversion markers. In 31 SMA patients, 45% of haplotypes showed varying *SMN1* to *SMN2* gene conversion breakpoints, serving as direct evidence of gene conversion as a common genetic characteristic in SMA and prompting further investigation into gene conversion markers as disease modifiers. Our findings illustrate that both methodological advances and the analysis of patient samples are required to advance our understanding of complex genetic loci and address critical clinical challenges.

## Introduction

The *SMN* locus on chromosome 5q, containing the *SMN1* and *SMN2* genes, consists of ∼2 Mb complex segmental duplications of a highly repetitive nature^1^. Homozygous loss-of-function of *SMN1* causes spinal muscular atrophy (SMA), a severe neuromuscular disease^2^. *SMN1* paralog *SMN2* has a paralogous sequence variant (PSV) in exon 7, causing most mRNA to be alternatively spliced and translated into an unstable, truncated protein (SMNΔ7)^3,4^. However, *SMN2* splicing also produces limited full-length mRNA that is translated into functional SMN protein, sufficient for survival but causing SMA in the absence of *SMN1*^5^. Although SMA is classified as a rare disease, its incidence of between 1:6,000-8,000 newborns suggest at any moment there may be up to 200,000 prevalent cases worldwide^6^. The severity of SMA ranges from prenatal onset and neonatal death (SMA type 0) to adult onset and a normal life expectancy (SMA type 4)^7^.

Loss of *SMN1* is hypothesized to be caused by homozygous deletion in severe types of SMA or gene conversion of *SMN1* to *SMN2* in comparatively milder types of SMA^8^. However, there is no direct evidence supporting this hypothesis and the frequency of such events remains undetermined. The total number of *SMN2* copies in the diploid human genome is highly variable and inversely correlates with the severity of SMA: a higher *SMN2* copy number leads to higher SMN protein levels and a relatively milder disease phenotype^9,10^. Yet, up to 40% of patients have a discordant disease phenotype relative to their *SMN2* copy number^9,11^. Earlier studies have identified rare *SMN2* sequence variants – such as c.859G>C and c.835-44A>G – as explanation for discordancy^12,13^. Copy number variation of surrounding genes such as *GTF2H2*, *(pseudo)NAIP* and *SERF1A/B*, has been associated with SMA disease severity in some reports, but this association remains inconclusive^14^. Moreover, fifteen PSVs have been described that can discriminate between *SMN1* and *SMN2* sequences^4,15,16^. These PSVs have been found to occur in different combinations as part of *SMN* hybrid genes and suggest the presence of extensively varying *SMN* haplotypes. However, the repetitive nature of the *SMN* locus represents a challenge for bioinformatic analysis from short-read sequencing data, complicating the identification of explanations for genotype-phenotype discordances in SMA^14^.

Recently, advancements in long-read sequencing have led to the assembly of novel telomere-to-telomere, pangenome reference sequences^17,18^. This fills many gaps in our knowledge of the human genome^18^ and includes the assembly of several novel *SMN* locus alleles^17,19^. However, these studies also highlight the complexity and diversity of genetic variation in this locus. After long-read sequencing, 65% of *SMN* alleles remained unresolved and the composition of resolved alleles varied extensively in gene copy number and orientation^17,19^. A recent PacBio HiFi long-read sequencing study on 438 samples, of which 11 known SMA carrier samples and one known SMA patient sample, identified a stretch of SNVs downstream of *SMN1* and *SMN2* that may be used to further characterize these haplotypes, but limited sequencing read length hampered analysis of larger flanks^20^. Many fundamental questions, including copy-specific *SMN2* sequence variation, the existence and frequency of *SMN1* to *SMN2* gene conversion events, the location and genetic environment of *SMN2* copies, and differences in composition of the *SMN* locus between healthy controls and patients with SMA, are yet to be answered. This shows that, despite using advanced techniques, the *SMN* locus remains one of the most complex in the human genome.

In addition to enhancing our understanding of complex loci in the human genome, several clinically relevant issues in SMA urgently need to be addressed. First, the incomplete correlation between SMA severity and *SMN2* copy number makes patient and family counseling challenging as disease severity cannot be precisely predicted^7,9^. Second, three gene-targeting therapies for SMA have been approved, two of which are *SMN2* splice-modifying treatments. There is significant variability in patient responses to these drugs^21,22^. The underlying causes of this variability are unknown, and individual treatment outcomes remain unpredictable. It is unclear how personal genetic variants influence treatment outcomes, despite the direct interaction of splice-modifying drugs with *SMN2* pre-mRNA. This underscores the need for an improved understanding of the composition of and genetic variation in the *SMN* locus, also from a clinical perspective^23^.

To address these issues, a more detailed study of the *SMN* locus in patients with SMA, supported by technological advances, is required. Here, we first developed HapSMA, a method to perform polyploid phasing of ∼173 kb of the *SMN* locus to enable copy-specific analysis of *SMN* and its surrounding genes, and performed long-read, targeted nanopore sequencing of the *SMN* locus of patients with SMA. Using HapSMA on publicly available healthy control data, we identified genetic variants that characterized the typical genetic environment of *SMN1* and *SMN2*, that could be used as markers for gene conversion. Using these variants in SMA patients, we identified highly variable haplotypes with varying *SMN1/2* gene conversion breakpoints in 45% of *SMN2* haplotypes. In summary, we here provide direct evidence of gene conversion as a common genetic characteristic in SMA and illustrate that both methodological advances and the analysis of patient samples are required to advance our understanding of complex genetic loci.

## Results

### Copy-specific analysis of *SMN* and surrounding genes by HapSMA: targeted mapping and polyploid phasing of nanopore sequencing reads

We first developed HapSMA, a method to enable sequence analysis of individual *SMN* copies and flanking regions. The *SMN* locus contains many segmental duplications, complicating mapping of both short-read and long-read sequencing data. To determine the extent and location of these duplications, we performed segmental duplication analysis on chromosome 5 of the telomere-to-telomere CHM13 (T2T-CHM13) reference genome (**Figure 1A**, **Figure S1A**). This analysis identified a ∼173 kb region, including *SMN2* and flanking sequences, of high similarity to *SMN1* and flanking sequences. This region was masked to prevent ambiguous mapping and to direct mapping of reads to *SMN1* and flanking region (∼173 kb), from here on called the phasing region of interest (ROI) (**Fig. 1B**). This allowed us to directly compare all *SMN1-* and *SMN2-*derived sequencing reads and identify variants that could reliably distinguish both genes and their environments. We performed nanopore sequencing of high molecular weight (HMW) DNA of 31 SMA patients (**Table 1**) and used adaptive sampling to enrich for a 30 Mb region surrounding the *SMN* locus (see **Materials & methods**). Whole-genome nanopore sequencing data of 25 healthy controls was available from the 1000 Genomes (1000G) project^24^. Applying our HapSMA bioinformatic workflow, sequencing reads were mapped to the masked T2T-CHM13 reference genome and phased into two to six haplotypes depending on total *SMN1/2* copy number as determined by MLPA^20,25^ (**Table S1**; **Fig. 1C**). An example of haplotype phasing across *SMN* and surrounding genes for one sample is shown in **Fig. 1D**. Median read depth on the 30 Mb region surrounding the *SMN* locus was 35.2x for 1000G and 22.3x for SMA (**Fig. 1E**). Read length N50 was similar between 1000G (52.5 kb) and SMA (48.6 kb) samples (**Fig. 1F**). Percentage of read depth on the phasing ROI that was successfully phased into haplotypes did not differ between 1000G (median = 77.5%) and SMA datasets (median = 74.9%; **Fig. 1G**). Minimum read depth for calling variants was set at 3x. Median per-haplotype coverage of the phasing ROI with a minimum read depth of 3x was 97.2% for 1000G and 79.6% for SMA (**Fig. 1H**), which was associated with total read depth per sample (**Fig. S1B**). Using HapSMA, we identified a median of 548 variants per sample (range 162-1149). In total, we identified 3908 unique variants, of which 2923 SNVs and 985 insertions/deletions (INDELs). To assess the performance of our method, we determined SNVs within the *SMN* gene using long-range PCR and Illumina sequencing in 29 of our SMA patients^15^, and found a high level of concordance between both methods (92%, **Fig. S1C**). Thus, haplotype phasing and copy-specific analysis of *SMN* genes including large flanks can be achieved with nanopore sequencing and HapSMA.

**Figure 1:**
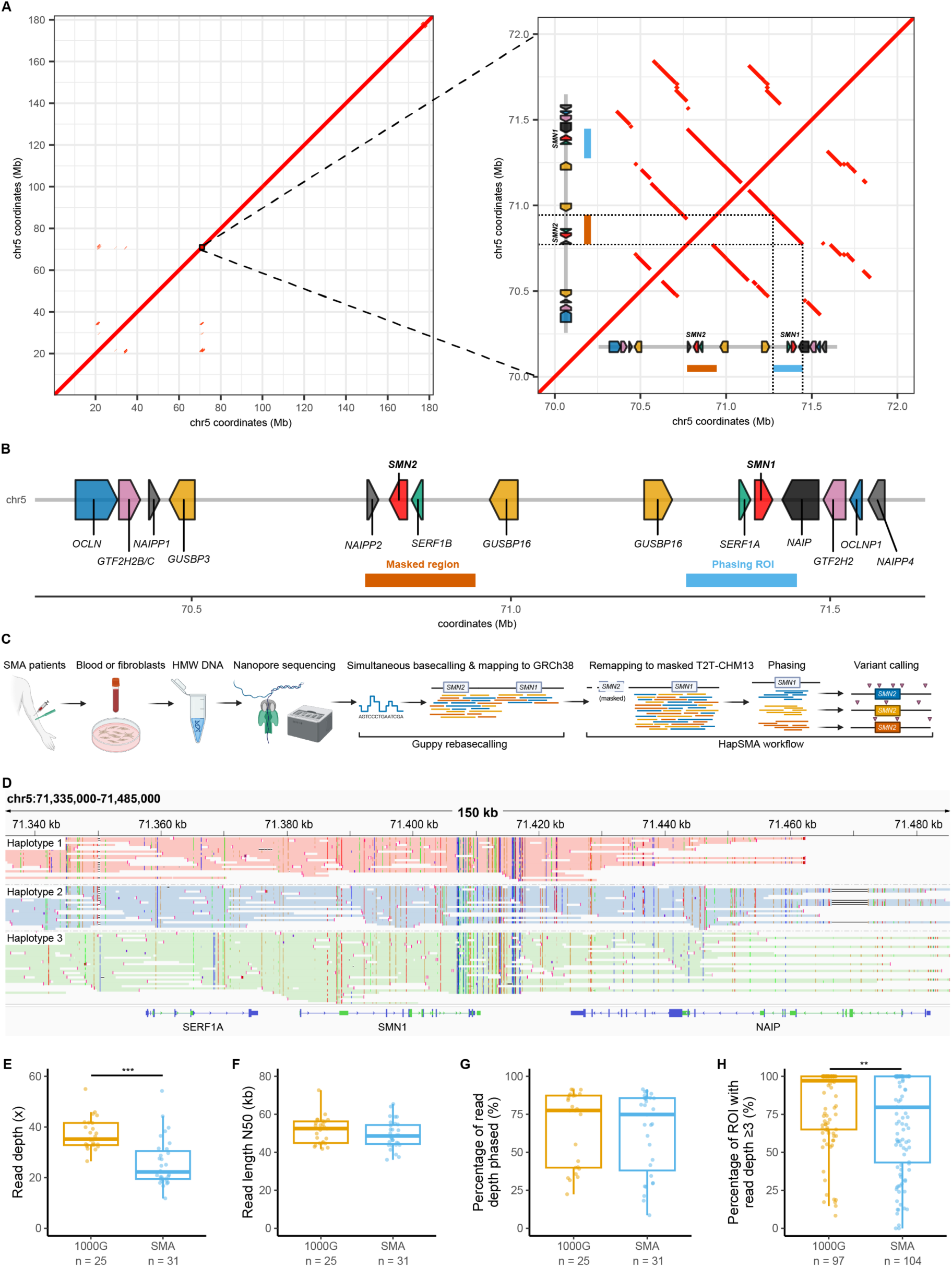
Copy-specific analysis of *SMN* and surrounding genes by targeted mapping and polyploid phasing of Nanopore sequencing reads **(A)** Dot plot resulting from segmental duplication analysis of T2T-CHM13 chromosome 5 against itself (left panel), zoomed in on the *SMN* locus (right panel). Red lines indicate segments of at least 95% similarity and at least 10 kb. The structure of the *SMN* locus as shown in B is shown at scale on both the x- and y-axis. **(B)** Structure of the *SMN* locus on the T2T-CHM13 reference genome. Genes are indicated by colored arrows. Genome coordinates chr5:70,772,138-70,944,284 were masked; and its segmental duplication counterpart chr5:71,274,893-71,447,410 was the main region of interest (ROI) for haplotype phasing. **(C)** Overview of sequencing and bioinformatics approaches in this study. Nanopore sequencing with adaptive sampling was performed on HMW DNA of SMA patients. Raw sequencing data was basecalled with the Guppy SUP model and mapped to GRCh38. Within the HapSMA workflow, reads were remapped to the masked T2T-CHM13 reference genome as indicated in (B). Polyploid variant calling and haplotype phasing was performed, followed by variant calling per haplotype. **(D)** Example of haplotype phasing of an SMA sample with 3 *SMN2* copies across *SMN1/2* and surrounding genes. Sequencing reads are colored by haplotype. Soft-clipping is not shown. **(E-H)** Overview of sequencing characteristics. (E) 1000G samples had median read depth of 35.2x, significantly higher than SMA samples (22.3x; Wilcoxon rank sum test, W = 664, p = 1.323e-06). (F) Read length N50 was not different between 1000G and SMA (52.5 kb and 48.6 kb, respectively; Wilcoxon rank sum test, W = 446, p = 0.3419). (G) Percentage of read depth that was phased into a haplotype was similar between 1000G (median = 77.5%) and SMA samples (median = 74.9%; Wilcoxon rank sum test, W = 424, p = 0.5559). (H) Percentage of phasing ROI with read depth 23 was higher for 1000G (median = 97.2%) than for SMA (median = 79.6%; Wilcoxon rank sum test, W = 6082, p = 0.008925). 1000G: 1000 Genomes; Mb: megabases

**Table 1:** Clinical characteristics and *SMN* copy number of SMA and 1000G samples. SMA patients were classified as concordant in case of 2x*SMN2* and Type 1, 3x*SMN2* and Type 2 or 3x*SMN2* and Type 3 (yellow); discordant (more severe) in case of 3x*SMN2* and Type 1, 4x*SMN2* and Type 2 or 5x*SMN2* and Type 3 (orange); discordant (less severe) in case of 2x*SMN2* and Type 3, 3x*SMN2* and Type 3 or 4x*SMN2* and Type 4 (green); 1000G: 1000 Genomes; F: female; M: male; NA: not applicable; MLPA: multiplex ligation-dependent probe amplification. ^a^Including patient with *SMN1* with c.542A>G mutation. ^b^Including patient with *SMN1* with deletion of exon 1-6.

|  | Total SMA | SMA Type 1 | SMA Type 2 | SMA Type 3 | SMA Type 4 | 1000G |
| --- | --- | --- | --- | --- | --- | --- |
| Sex (F:M) | 13:18 | 3:2 | 5:8 | 5:5 | 0:2 | 18:7 |
| Median age at onset in years (range) | 1.0 (0.0-24.5) | 0.4 (0-0.5) | 0.9 (0.5-1.5) | 7 (1-15.5) | 22.5 (20.5-24.5) | NA |
| Median age at sampling in years (range) | 9.7 (0.4-70.1) | 4.6 (0.4-41.5) | 8.0 (2.1-57.5) | 18.1 (5.2-63.3) | 60.3 (50.4-70.1) | NA |
| **SMN2 copy number (MLPA)** |  |  |  |  |  |  |
| 0x SMN2 |  |  |  |  |  | 2 |
| 1x SMN2 |  |  |  |  |  | 11 |
| 2x SMN2 | 3 | 2 |  | 1<sup>a</sup> |  | 11 |
| 3x SMN2 | 16 | 3 | 10<sup>b</sup> | 3 |  | 1 |
| 4x SMN2 | 11 |  | 3 | 5 | 2 |  |
| 5x SMN2 | 1 |  |  | 1 |  |  |
| **SMN1 copy number (MLPA)** |  |  |  |  |  |  |
| 0x SMN1 | 29 | 5 | 12 | 9 | 2 |  |
| 1x SMN1 | 2 |  | 1<sup>b</sup> | 1<sup>a</sup> |  |  |
| 2x SMN1 |  |  |  |  |  | 17 |
| 3x SMN1 |  |  |  |  |  | 6 |
| 4x SMN1 |  |  |  |  |  | 2 |

### Specific SNVs and *NAIP* variants mark *SMN1* and *SMN2* environments in healthy controls

Our current knowledge of *SMN1-* or *SMN2*-specific variants is mostly limited to the presence of 15 intragenic PSVs^4,15,16^. To expand the range of variants capable of differentiating *SMN1* and *SMN2* and their broader genetic environments, we first used publicly available Nanopore sequencing data of 25 samples from the 1000G project^24^ of healthy controls (**Table 1**) and applied HapSMA to determine 97 copy-specific *SMN1* and *SMN2* haplotypes (**Fig. 2A**). Haplotypes were classified as *SMN1* or *SMN2* based on PSV13 (c.840C>T variant). Genetic variants were classified as *SMN2*-specific, when at least 90% of *SMN2* haplotypes and a maximum of 20% of *SMN1* haplotypes contained the variant. At these cutoffs, all 15 known PSVs were detected as *SMN2*-specific variants, in contrast to stricter cutoffs of 90% and 10%, in which case PSV1 and PSV5 would be excluded. In addition, we identified four SNVs upstream of *SMN1/2* and *SERF1A/B*, and 23 downstream SNVs (**Fig. 2A**), from here on called *SMN2* environment SNVs. Twenty-one of these downstream SNVs have been reported before using PacBio sequencing and the bioinformatic tool Paraphase^20^, confirming the validity of our results. No *SMN1*-specific variants were found using these criteria; the presence of the reference allele on an *SMN2*-specific variant position was thus defined as an *SMN1* environment SNV.

**Figure 2:**
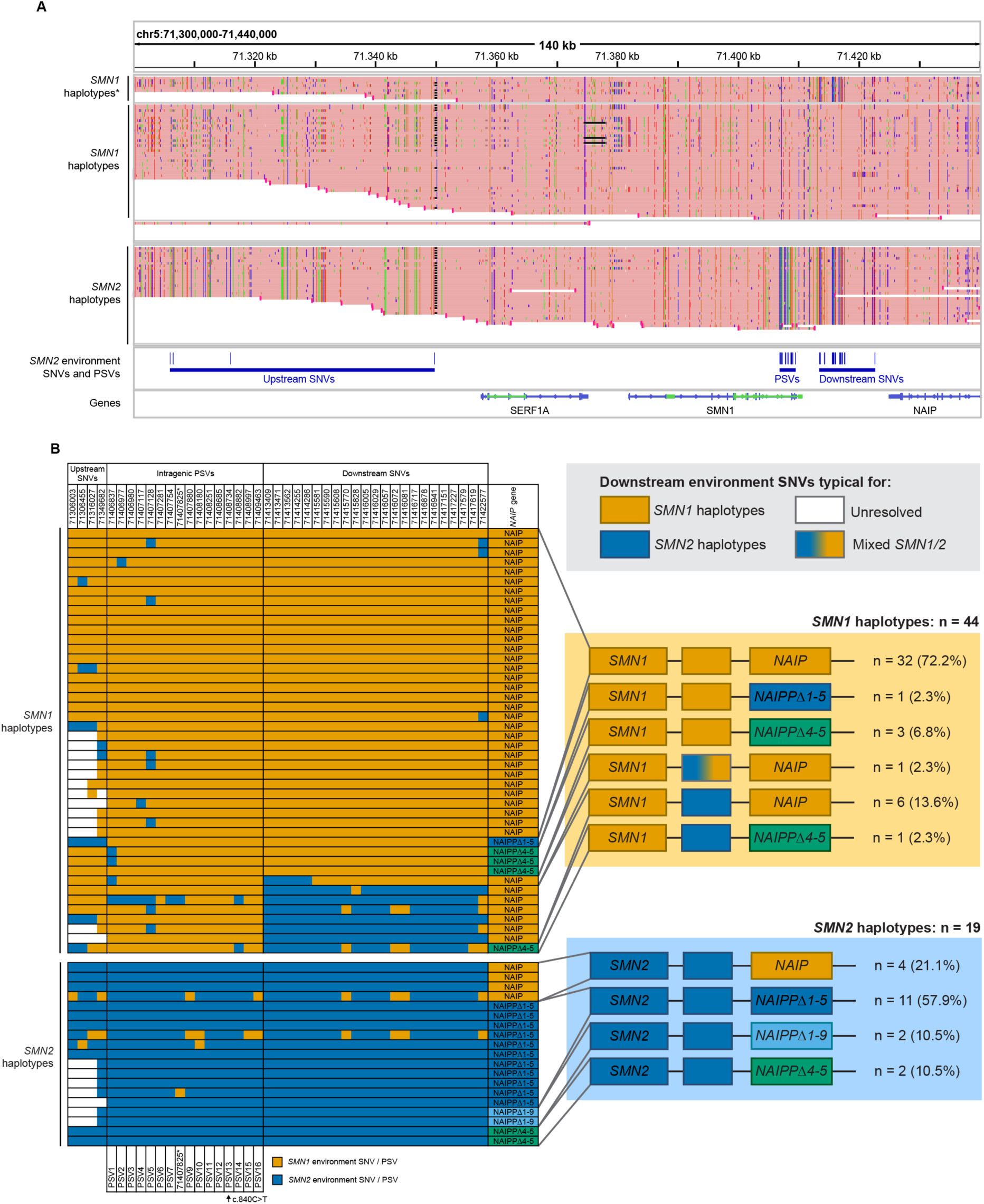
In healthy controls, the upstream and downstream environments of *SMN2* are characterized by specific SNVs and *NAIP* variants **(A)** IGV overview of *SMN1* and *SMN2* haplotypes (divided based on the c.840C>T variant in exon 7 (see **Materials and Methods**)) from 1000G healthy control samples, mapped to the T2T-CHM13 masked reference genome. Each ‘read’ represents one haplotype from one sample. *SMN2*-specific variants (present in ≥90% of *SMN2* haplotypes and ≥20% of *SMN1* haplotypes), indicating an *SMN2* environment, are indicated by blue lines in the lower panel, including PSVs, upstream SNVs and downstream SNVs. \**SMN1* haplotypes with *SMN2*-specific downstream SNVs. **(B)** Schematic representation of PSVs, *SMN1/2* environment SNVs and presence of the *(pseudo)NAIP* gene per haplotype. Only haplotypes with complete phasing between the c.840C>T PSV (indicated with an arrow) and *(pseudo)NAIP* are shown. In the right panel, downstream haplotype frequencies are shown schematically. Downstream environment other than the ‘expected’ environment was called when 3 or more consecutive *SMN1/2* environment SNVs were present. Full-length *NAIP* was characterized as *SMN1* environment, whereas truncated *NAIPPΔ1-5* or *NAIPPΔ1-9* was characterized as *SMN2* environment^20^. *NAIPPΔ4-5* was not characterized as either environment. *The 5 bp insertion at position 71407825 was not included as PSV. *NAIPP*: *pseudoNAIP*; PSV: paralogous sequence variant; SNV: single nucleotide variant.

Next, we investigated the presence of the *(pseudo)NAIP* gene downstream of *SMN* in each haplotype. In most previously published haploid reference alleles (with one copy *SMN1* and one copy *SMN2*), *NAIP* is adjacent to *SMN1*, and a truncated *pseudoNAIP* is adjacent to *SMN2*^20^. Therefore, we defined full-length *NAIP* as *SMN1* environment. In addition to full-length *NAIP*, we observed different types of truncated *pseudoNAIP* (*NAIPP*) genes downstream of *SMN1/2*: *NAIPPΔ4-5* lacking exon 4-5, *NAIPPΔ1-5* lacking exon 1-5 and *NAIPPΔ1-9* lacking exon 1-9 (**Fig. S2**). We defined *NAIPPΔ1-5* and *NAIPPΔ1-9* as *SMN2* environment, but as *NAIPPΔ4-5* was not clearly enriched in either *SMN1* or *SMN2* haplotypes, it was not defined as specific for either environment. For 63 of the 97 1000G haplotypes (64.9%), the *(pseudo)NAIP* gene was phased successfully. These haplotypes are shown in **Fig. 2B**; with a more extensive list in **Table S1**. In 32 of 44 *SMN1* haplotypes (72.7%), only elements of a downstream *SMN1* environment (downstream *SMN1* environment SNVs and full-length *NAIP*) were present, whereas eight haplotypes (18.2%) contained an element of downstream *SMN2* environment (downstream *SMN2* environment SNVs, *NAIPPΔ1-5*, *NAIPPΔ1-9* or a combination). Four *SMN1* haplotypes (9.1%) contained *NAIPPΔ4-5*. Thirteen (68.4%) *SMN2* haplotypes had a full downstream *SMN2* environment, and four (21.1%) a downstream *SMN1* environment. Two (10.5%) contained downstream *NAIPPΔ4-5*. An upstream environment different from the expected environment was rare: three (9.7%) of 31 completely phased *SMN1* haplotypes had an upstream *SMN2* environment, and zero out of 12 completely phased *SMN2* haplotypes had an upstream *SMN1* environment (**Fig. 2B**). In summary, our results show typical genomic environments for *SMN1* and *SMN2* in healthy controls, based on the analysis of SNVs and *NAIP* variants.

### Markers for gene conversion are abundant in SMA patients and the underlying recombination events can occur at different breakpoints downstream of *SMN2*

Next, we determined the presence of the identified markers of *SMN1* and *SMN2* environments in SMA patient haplotypes (**Fig. 3A**). For 53 of 104 SMA haplotypes (51.0%), the *(pseudo)NAIP* gene was phased completely. We included two patients with pathogenic variants in *SMN1* and found that, as expected, both *SMN1* haplotypes had a downstream *SMN1* environment. Amongst *SMN2* haplotypes, different types of downstream *SMN1* environments were present: 1) all downstream *SMN1* environment SNVs and *NAIP*, 2) three or more consecutive downstream *SMN1* environment SNVs and *NAIP*, or 3) fewer than three consecutive downstream *SMN1* environment SNVs and *NAIP*. This indicates that the recombination events underlying gene conversion in SMA occur at different genomic locations (**Fig. 3B**). Out of 39 non-hybrid *SMN2* haplotypes, 21 haplotypes (53.8%) had a full downstream *SMN2* environment, 15 haplotypes (38.5%) had a full or partial downstream *SMN1* environment, and three haplotypes (7.7%) had an ambiguous environment characterized by presence of *NAIPPΔ4-5*. In contrast to controls, an upstream environment different from the expected environment was common, as it was present in 11 out of 24 fully resolved non-hybrid *SMN2* haplotypes (45.8%).

**Figure 3:**
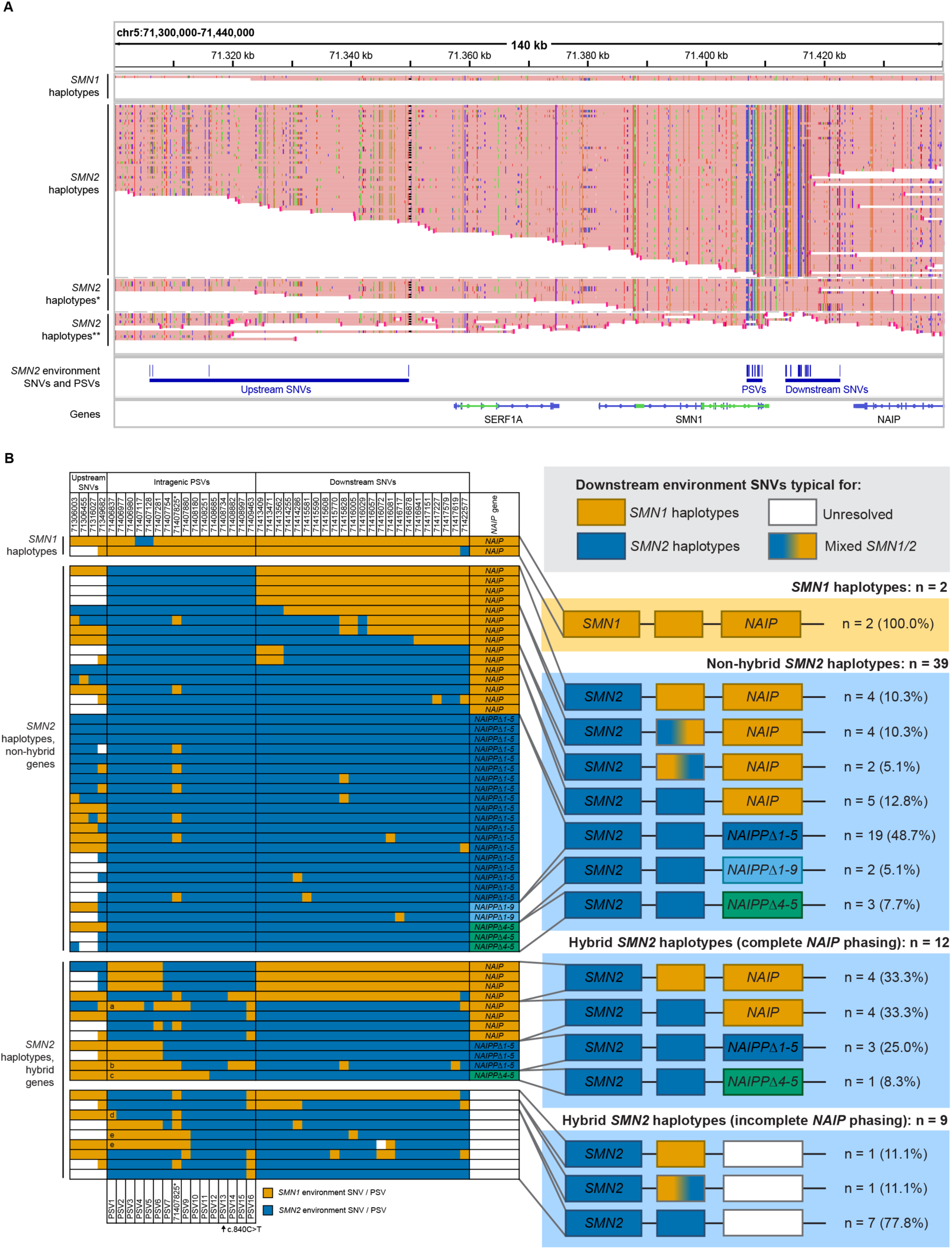
Markers of *SMN1* environment are abundant and highly variable in *SMN2* haplotypes of SMA patients **(A)** IGV overview of *SMN1* and *SMN2* haplotypes (divided based on the c.840C>T variant in exon 7 (see **Materials and Methods**)) from SMA patients, mapped to the T2T-CHM13 masked reference genome. Each ‘read’ represents one haplotype from one sample. *SMN2*-specific variants as determined in Figure 2A, indicating an *SMN2* environment, are indicated by blue lines in the lower panel, including PSVs, upstream SNVs and downstream SNVs. \**SMN2* haplotypes with a downstream *SMN1* environment. \*\**SMN2* haplotypes with an incompletely resolved downstream *SMN1/2* environment. **(B)** Schematic representation of PSVs, *SMN1/2* environment SNVs and presence of the *(pseudo)NAIP* gene per haplotype. Of non-hybrid *SMN2* haplotypes, only haplotypes with complete phasing between the c.840C>T PSV (indicated with an arrow) and *(pseudo)NAIP* are shown. In the two lower left panels, haplotypes with hybrid SMN2 genes are shown, of which five hybrid structures are novel: PSV1-4, 6-9 and 16 (a); PSV1-7 and 14-16 (b); PSV1-11 (c); PSV1 (d); PSV1-9 (e). In the right panel, downstream haplotype frequencies are shown schematically. Downstream environment other than the ‘expected’ environment was called when 3 or more consecutive *SMN1/2* environment SNVs were present. Full-length *NAIP* was characterized as *SMN1* environment, whereas truncated *NAIPPΔ1-5* or *NAIPPΔ1-9* was characterized as *SMN2* environment^20^. *NAIPPΔ4-5* was not characterized as either environment. The percentage of downstream *SMN1* environment in hybrid haplotypes (66.7%) was not significantly higher than in non-hybrid *SMN2* haplotypes (38.5%; Fisher’s exact test, p = 0.107). *The 5 bp insertion at position 71407825 was not included as PSV. *NAIPP*: *pseudoNAIP*; PSV: paralogous sequence variant; SNV: single nucleotide variant.

Previously, hybrid genes have been noted as markers of gene conversion events^26^, suggesting they are more likely to have a downstream *SMN1* environment. Here, we found 10 different *SMN* hybrid gene structures across 21 haplotypes, five of which were novel (**Fig. 3B**, two lower left panels). From 12 completely resolved hybrid *SMN2* haplotypes, three (25.0%) contained a downstream *SMN2* environment, eight (66.7%) contained a downstream *SMN1* environment, and one (8.3%) contained an unclear (*NAIPPΔ4-5*) environment. The percentage of downstream *SMN1* environment in hybrid haplotypes (66.7%) was not significantly higher than in non-hybrid *SMN2* haplotypes (38.5%; Fisher’s exact test, p = 0.107). However, all hybrid haplotypes with unresolved or *SMN2* downstream environments had only upstream *SMN1* environment SNVs (100%, n=5). This supports the theory that hybrid *SMN2* genes likely arose from gene conversion. We next hypothesized that the upstream or downstream environments of *SMN2* may affect its activity through regulatory elements such as enhancers. Exploratory analyses comparing *SMN* gene environments to SMA type, and *SMN2-Δ7/FL* mRNA and SMN protein expression illustrate substantial variability that warrants further study in a larger cohort (**Fig. S3**). In summary, we provide evidence for gene conversion in 45% of phased *SMN2* haplotypes and show that the underlying recombination events can occur at different breakpoints downstream of *SMN2*.

### Copy-specific sequence and structural variants are common in patients with SMA

Because HapSMA allows for copy-specific analysis, we next assessed the presence of copy-specific exonic and structural variants. We detected several types of exonic variants in SMA patients: three missense, two synonymous and two untranslated region (UTR) variants (**Table 2**). The pathogenic missense variant c.542A>G (p.Asp181Gly), that was reported previously^11^, was confirmed to be present in exon 4 of *SMN1* in one patient. It is predicted to create a new splice-donor site within exon 4 of *SMN1* leading to a truncated transcript by introducing a preliminary stop codon^11^. The mild phenotype of this patient (SMA type 3a) with respect to their *SMN2* copy number (two) suggests residual activity of this *SMN1* copy. Indeed, a small amount of *SMN1-FL* mRNA could still be detected (**Fig. S4A**). Other known positive modifying variants, such as c.859G>C and c.835-44A>G^12,13^, were not detected in this patient. Two missense variants in *SMN2*, c.77G>A present in two patients and c.593C>T in one patient, were predicted to be pathogenic and thus candidate negative modifiers (**Supplementary Note 1**). We found no clear link between the abovementioned exonic variants and *SMN* mRNA or SMN protein level in patients’ derived fibroblasts with and without the variant (**Fig. S4B-D**).

**Table 2:** Exonic variants in *SMN1* and *SMN2* identified by nanopore sequencing. Annotation and variant calls were based on *SMN1* transcript ENST00000380707.9.

| Gene with variant | HGVSc | HGVSp | Variant consequence | SIFT | PolyPhen | Alpha Missense | CADD PHRED | # of haplotypes | # of patients | Patient phenotypes | Reported before in SMA |
| --- | --- | --- | --- | --- | --- | --- | --- | --- | --- | --- | --- |
| *SMN1* * | c.542 A>G | p.Asp181Gly | missense | deleterious | benign | likely benign | 22.2 | 1/89 | 1 | Less severe | Wadman et al., 2020 |
| *SMN1* * | c.835-96A>G | - | intron | - | - | - | 14.87 | 1/93 | 1 | Less severe | This study |
| *SMN2* | c.-14 C>T | - | 5' UTR | - | - | - | 4.697 | 1/84 | 1 | Concordant | Wadman et al., 2020 |
| *SMN2* | c.77 G>A | p.Gly26Asp | missense | deleterious | possibly damaging | likely pathogenic | 24.5 | 2/85 | 2 | More severe, concordant | This study |
| *SMN2* | c.81+45C>T | - | intron | - | - | - | 5.642 | 1/84 | 1 | Concordant | Wadman et al., 2020 |
| *SMN2* | c.84 C>T | p.Ser28Ser | splice region, synonymous | - | - | - | 11.69 | 3/88 | 3 | More severe, concordant, less severe | Ruhno et al., 2019; Wadman et al., 2020 |
| *SMN1* */ *SMN2* | c.462 A>G | p.Gln154Gln | synonymous | - | - | - | 8.282 | 55/89 | 24 | - | Ruhno et al., 2019; Wadman et al., 2020 |
| *SMN2* | c.593 C>T | p.Pro198Leu | missense | deleterious | probably damaging | likely benign | 19.94 | 2/89 | 1 | More severe | This study |
| *SMN2* | c.723+59G>C | - | intron | - | - | - | 5.842 | 4/90 | 4 | Less severe (2), concordant (1), more severe (1) | Wadman et al., 2020 |
| *SMN2* | c.724-54C>T | - | intron | - | - | - | 7.545 | 10/92 | 8 | Less severe (2), concordant (6) | Wadman et al., 2020 |
| *SMN2* | c.*58 G>A | - | 3' UTR | - | - | - | 10.33 | 1/94 | 1 | Concordant | This study |

\**SMN1* with known pathogenic variant

In addition to small sequence variants, we also identified several structural variants in *SMN* and its surrounding genes. First, in one SMA patient we detected four copies of *SMN* exon 7-8 -one of which derived from *SMN1* - and three copies of *SMN* exon 1-6, indicating either an exon 1-6 deletion or *SMN1* exon 7-8 insertion. Either situation indicates an incomplete *SMN1* copy containing only exon 7 and 8, which corresponded with the absence of detectable *SMN1* mRNA in this patient (**Fig. S4A**). The *SMNΔ7-8* 6.3 kb deletion^25^ – relatively common in some populations but not in the Netherlands – was found in one 1000G haplotype (**Fig. S5**) but not in any of our SMA patients. Further structural variants were identified in *(pseudo)NAIP* (**Fig. S6**), and *SERF1A/B* (**Fig. S7)**. Unfortunately, an incomplete understanding of the functionality of *SERF1* and *NAIP* (pseudo)genes, however, prevents us from further speculating on the functional relevance of these structural variants. In summary, HapSMA allows for detection of sequence and structural variants in a haplotype-aware manner, which provides useful information about the localization of different variants on the same or different copies of *SMN* and surrounding genes, improving opportunities to interpret the possible functional effect of *SMN* variants.

## Discussion

A complete understanding of genetic variation in the complex *SMN* locus requires both methodological advances and its study in the genomes of SMA patients. Therefore, we performed copy-specific haplotype phasing of the ∼173 kb genomic environment surrounding *SMN* gene copies in a large cohort of SMA patients using targeted long-read sequencing. We identified upstream and downstream *SMN1* environment SNVs serving as markers of gene conversion, and the downstream presence of varying *NAIP* (pseudo)genes as markers of genomic location. These markers provide direct evidence of *SMN1* to *SMN2* gene conversion as a common genetic characteristic of SMA. We found that broad phasing including the *NAIP* gene allowed for a more complete view of *SMN* locus variation, increasing the number of identified downstream environment types from four to seven compared to when only downstream SNVs would have been considered^20^. Moreover, both the number of different haplotypes and the genetic variation in *SMN2* haplotypes was larger in patients with SMA, highlighting the importance of inclusion of SMA patients when investigating the *SMN* locus.

By examining *SMN1* environment SNVs in *SMN2* haplotypes, we determined that the recombination events underlying gene conversion can happen at different breakpoints between *SMN* and *NAIP*. Occasionally, more than one recombination breakpoint is present within one haplotype. The high variability of *SMN2* haplotypes in SMA is a confirmation of the high variability in the *SMN* locus between individuals, as reported before^19^. Previously, *SMN2* hybrid genes have been used as markers for gene conversion^26^, mostly based on PSV16 in exon 8. In this study, we show that upstream or downstream markers of *SMN1* environment are also present in all resolved haplotypes containing a hybrid *SMN2* gene. This strengthens the theory that *SMN2* hybrid genes likely arose from gene conversion. In addition, we also identified hybrid gene structures in patients without the PSV16 variant, indicating that absence of this variant cannot be used to exclude the presence of gene hybrids on its own. Twenty-one of 23 downstream *SMN2* environment SNVs identified in our study were previously also identified in *SMN1* haplotypes by PacBio HiFi sequencing^20^. Interestingly, similar to our findings, *SMN2* haplotypes with downstream *SMN1* environment SNVs were not detected in non-SMA individuals^20^.

We showed that different *(pseudo)NAIP* genes can be present downstream of *SMN1/2*. A previous study showed that on ‘normal’ alleles with one copy of *SMN1* and one copy of *SMN2*, the vast majority of *SMN1* copies were upstream of full-length *NAIP* and *SMN2* copies were upstream of truncated *pseudoNAIP*^20^. In 1000G control data, we similarly observed that *SMN1* was most often followed by *NAIP*, whereas *SMN2* was most often followed by truncated *NAIPPΔ1-5* or *NAIPPΔ1-9*. In contrast, *NAIPPΔ4-5* was found downstream of both *SMN1* and *SMN2* in similar frequencies and was therefore not categorized as specific *SMN1* or *SMN2* haplotypes. When assuming inverse gene orientation for *SMN1* and *SMN2* as on the T2T-CHM13 reference genome, this could happen through fusion deletion between *NAIPP1* and *NAIPP2* for *SMN2* haplotypes or inversion for *SMN1* haplotypes. Therefore, we hypothesize that the presence of *NAIPPΔ4-5* downstream of *SMN1/2*, which occurs in ∼8% of *SMN2* copies, indicates its localization next to *OCLN* and *GTF2H2C*. However, *de novo* assembly of the complete *SMN* locus in a sufficiently large number of genomes is required to confirm this. No *SMN1/2*-environment-specific variants were found in the *SERF1A/B* gene, suggesting that *SERF1A* and *SERF1B* are identical. The consistent co-phasing of *SERF1A/B* with *SMN* and absence of clear structural variation breakpoints confirm that most genomic rearrangements causing deletions associated with SMA include the *SERF1A/B* gene^20,27^.

Our newly identified markers of gene conversion and genomic location provide a foundation for further studies into their possible prognostic potential in larger cohorts. One hypothesis is that variation in genetic environment of *SMN* may affect RNA and protein levels by influencing local regulatory elements such as enhancers or DNA methylation, which is supported by exploratory analyses of SMN protein and mRNA analyses included in our study. However, data on chromatin accessibility (H3K27ac ChIP-Seq and ATAC-seq), 3D DNA interactions (Hi-C) and DNA methylation are currently limited within the *SMN* locus, as this type of data is largely based on short-read sequencing and thus, does not map well on the *SMN* locus^28–30^ or has not yet been studied in significant detail^31^. However, long-read methods for chromatin accessibility and interactions are emerging, and could prove useful in identifying regulatory regions within the *SMN* locus^32–34^. Similarly, methods to obtain information on base modifications such as methylation from long-read sequencing reads are increasingly sensitive and reliable^35,36^, opening possibilities to obtain this information concurrently with sequence and structural variation. These developments may also be useful in estimating the potential functional effects of intergenic structural variants such as the Alu insertions and deletions identified upstream of *SERF1A/B*. Furthermore, HapSMA enables the identification of the specific *SMN* copy on which a pathogenic variant is present, which supports the prediction of remaining SMN functionality in case of multiple pathogenic variants.

While our understanding of the complexity of the *SMN* locus has advanced considerably, it remains one of the most elusive regions in the human genome. This study, along with advancements in technology and the increasing cost-effectiveness of long-read sequencing, holds the promise of obtaining a more nuanced understanding of this locus and its relationship to SMA. Our findings reveal a heightened complexity in the *SMN* locus of SMA patients when compared to healthy controls, underscoring the necessity of including SMA patients in analyses of *SMN* locus composition and variation. The broad heterogeneity we observed suggests that extensive *de novo* assembly of both control and SMA patient samples will be essential to achieve a comprehensive understanding of the *SMN* locus. This will not only be vital for enhancing our basic understanding of complex genetic loci but also for addressing critical clinical challenges in SMA.

## Acknowledgements and funding

This work was supported by grants from Stichting Spieren voor Spieren (to WLvdP), the European Union’s Horizon 2020 Research and Innovation Program under the Marie Skłodowska-Curie grant (H2020 Marie Skłodowska-Curie Actions) agreement no. 956185 (SMABEYOND ITN to WLvdP, EJNG, EFT) and Prinses Beatrix Spierfonds (W.OB21-01 to EJNG). This project has received funding from the European Research Council (ERC) under the European Union’s Horizon 2020 research and innovation programme (grant agreement n° 772376 - EScORIAL). This work was partially funded by grants to EFT from Biogen (ESP-SMG-17-11256), Roche, GaliciAME and the Spanish Instituto de Salud Carlos III, Fondo de Investigaciones Sanitarias and co-funded with ERDF funds (grant no. FIS PI18/000687). We acknowledge the Utrecht Sequencing Facility (USEQ) for providing sequencing service and data. USEQ is subsidized by the University Medical Center Utrecht and The Netherlands X-omics Initiative (NWO project 184.034.019). We thank B.P.C. Koeleman for critically reading this manuscript and providing extensive feedback.

## Author contributions

Conceptualization, MMZ, MGE, DG, EJNG, GWvH; experimental studies, MMZ, IS, LBP, MCR, IRJG, JK; bioinformatics, MMZ, MGE, DG, JvdS, CV, RS; clinical data & patient material, FA, RIW, WLP; writing–original draft, MMZ, EJNG, GWvH; writing–review, all; resources, EFT, WLvdP, EJNG; supervision, WLvdP, EJNG, GWvH; funding acquisition, EJNG, GWvH, WLvdP, EFT.

## Methods

### Study population

31 SMA patients with a variety of *SMN* copy numbers and SMA types (**Table 1**) were included from our single-center prevalence cohort study in the Netherlands. The study protocol (09307/NL29692.041.09) was approved by the Medical Ethical Committee of the University Medical Center Utrecht and registered at the Dutch registry for clinical studies and trials (https://www.ccmo.nl/). Written informed consent was obtained from all adult patients, and from patients and/or parents additionally in case of children younger than 18 years old. *SMN1*, *SMN2* and *NAIP* copy number was determined using multiplex ligation-dependent probe amplification (MLPA) (MRC Holland, SALSA MLPA Probemix P021 SMA Version B1) according to the manufacturer’s protocol (https://www.mrcholland.com/). Clinical SMA type was determined as described previously^37^. Patients were classified as concordant or discordant (less or more severe) based on deviation of clinical SMA type from MLPA-determined *SMN2* copy number; concordant being SMA type 1 with two copies of *SMN2*, SMA type 2 with three copies of *SMN2*, SMA type 3 with four copies of *SMN2*, and SMA type 4 with five copies of *SMN2*^7,9^. To represent the wide genotypic and phenotypic variety in the SMA population, we included patients with *SMN2* copy number ranging from two to five, and SMA type ranging from 1b to 4. Whole blood was obtained in EDTA blood tubes for DNA extraction and 3 mm dermal biopsies were obtained for generation of primary fibroblasts.

### Culture of primary fibroblasts

Patient-derived fibroblasts were cultured in Dulbecco’s modified Eagle’s medium (DMEM, Gibco, 41966-029) containing 4.5g/L D-glucose, L-glutamine and pyruvate, supplemented with 10% heat-inactivated fetal bovine serum (Cytvia, SH30073.03) and penicillin-streptomycin (Sigma-Aldrich, P0781), in a humidified 5% CO2 atmosphere, at 37°C. At a confluency of 70-80%, fibroblasts were enzymatically detached with Accutase (Sigma Aldrich, A6964) and frozen at -80°C as pellets for RNA or protein extraction, or resuspended in DMEM containing 20% FBS and 20% dimethyl sulfoxide (DMSO) for HMW DNA extraction.

### RNA and protein quantification using droplet digital PCR (ddPCR) and western blot

RNA was extracted from fibroblasts with the RNeasy mini kit (Qiagen, 74104) and total RNA was treated with DNaseI treatment (Thermo Scientific, EN0521). RNA concentration was quantified with a spectrophotometer (Nanodrop 2000, Thermo Scientific). cDNA was synthesized from 100 ng of RNA using the high-capacity cDNA reverse transcription kit (Applied Biosystems, 4368814). *SMN1-FL*, *SMN2-FL*, *SMN2Δ7* and *TBP* mRNA levels were quantified by ddPCR with primers and probes (Integrated DNA Technologies) in three technical replicates as described previously^11,38^. Expression levels of *SMN1-FL, SMN2-FL* and *SMN2Δ7* were normalized against *TBP* expression using QuantaSoft Software (Bio-Rad, 1864011). Protein was extracted from fibroblast pellets by homogenization in RIPA buffer (Thermo Scientific, 89900) with 1x protease inhibitor (Thermo scientific, 1861278), incubation on ice for 10 min and centrifugation at 4°C at 18,620x rcf for 10 min. Supernatant was collected and protein concentration was measured with the micro BCA protein assay kit (Thermo scientific, 23235). For SMN protein quantification, semi-quantitative western blotting was performed in technical triplicates as described previously^39^. SMN protein level, determined with mouse-anti-SMN primary antibody (BD Bioscience, 610647; 1:1000) and donkey-anti-mouse secondary antibody IRDye 800 (Licor, 926-32212; 1:2500), was normalized against Revert 700 total protein stain (Licor, 926-11011). Results were normalized to an internal standard (SMN level from HEK293 cell lysates) to enable reliable comparison of quantifications obtained from different membranes.

### HMW DNA extraction

For SMA patients, high molecular weight (HMW) DNA was extracted from fresh or frozen whole EDTA blood (n = 8), cultured primary dermal fibroblasts (n = 20) or both tissues (n = 3) using the Monarch® HMW DNA Extraction Kit for Cells & Blood (New England Biolabs (NEB), T3050L) with lysis agitation at 1400 rpm. For three samples, additional ultra-high molecular weight (UHMW) DNA was extracted from fibroblasts with an adapted protocol according to Ultra-Long Sequencing Kit (Oxford Nanopore Technologies (ONT), SQK-ULK001) guidelines. DNA concentration was measured using the Qubit™ dsDNA Quantification Broad Range Assay (Invitrogen, Q32853) and purity was determined on a spectrophotometer (Nanodrop 2000, Thermo Scientific).

### Segmental duplication analysis and masking

We used the recently published T2T-CHM13 genome^18^ as the reference genome for our analyses, as it has been shown to be more suitable for analyzing complex regions than previous reference genomes^40^. Segmental duplication analysis of T2T-CHM13 chromosome 5 against itself was performed with the nucmer command within MUMmer 3.23^41^. Nucmer output was filtered for segments with a sequence identity score of ≥95%, length of ≥10 kb and coordinates between 70 and 72 Mb, visualized in R (v4.4.0) and converted into a bed file for visualization in IGV. Each segment was assigned a unique identifier from 1a to 59a, with corresponding duplicated counterparts labeled as 1b to 59b. Based on this analysis, segments 20a and 22a (chr5:70,772,138-70,944,284) were masked, to direct mapping of *SMN1* and *SMN2* sequencing reads to segments 20b and 22b. We defined this as our phasing ROI (chr5:71,274,893-71,447,410).

### Alignment of NAIP and pseudoNAIP genes

*NAIP* and *pseudoNAIP (NAIPP)* sequences with 10 kb flanks were extracted from the T2T-CHM13 reference genome: *NAIP* (NCBI RefSeq NM_004536.3), CAT/Liftoff *NAIPP1-201*, CAT/Liftoff *NAIPP2-201*, NCBI RefSeq *NAIPP3* and NCBI RefSeq *NAIPP4*. All *NAIPP* genes were aligned with the full-length *NAIP* gene with the megablast setting in NCBI Nucleotide BLAST.

### Sequencing

Per sequencing run, three library preps with 1.3 µg HMW DNA each were made with the ligation sequencing kit (ONT, SQK-LSK109). For the UHMW samples, the Ultra-Long Sequencing Kit (ONT, SQK-ULK001) was used. The library preps were sequenced on a FLO-MIN106 flow cell on a GridION (ONT) with MinKNOW v21.02.5-22.12.5 and FAST basecalling for 72 hours, with a nuclease flush and reloading a new library prep every 24 hours. Adaptive sampling^42,43^ was used within MinKNOW, with a combined target FASTA file (see **Supplementary materials**) of the 30 Mb region surrounding the *SMN* locus (GRCh38 chr5:55,000,000-85,000,000) and six resolved alleles of the *SMN* locus downloaded from https://zenodo.org/records/5501736^19^. Long-range PCR and Illumina sequencing was performed as previously published^15^.

### Dataprocessing: rebasecalling and data download

For the SMA patients, rebasecalling of the raw sequencing data (FAST5 files) was performed using the SUP model in Guppy v6.1.2, including mapping to GRCh38.

Nanopore BAM files from the 1000 Genomes (1000G) dataset, mapped to GRCh38, were downloaded^24^ and 25 samples for which *SMN1/2* copy number was known^20,25^ (**Table 1**) were used in this study. Ethnic origins were England (n=7), Finland (n=1), Spain (n=3), China (n=4), Bangladesh (n=2), Nigeria (n=5), Gambia (n=1) and Sierra Leone (n=2). Reads mapped to chromosome 5 were extracted with samtools v1.17.

### Directed mapping, polyploid haplotype phasing and variant calling

BAM files from the SMA patients and 1000G samples were processed using the HapSMA workflow v1.0.0 (available on https://github.com/UMCUGenetics/HapSMA/) with option bam_remap (SMA data, using multiple BAM output files from Guppy) or bam_single_remap (1000G data, using a single BAM file). In short, sequencing reads were re-mapped to the masked T2T-CHM13 reference genome using minimap2 v2.26, and ploidy-aware variant calling was performed on the phasing ROI using GATK v4.2.1.0 based on *SMN1/2* total copy number determined by MLPA. The SNVs from the resulting VCFs were used for haplotype phasing with Whatshap v1.7. The BAM file was then split for each haplotag (HP) of the phaseset (PS) covering *SMN1* using sambamba v1.0.0, resulting in one BAM file per haplotype. Sequencing reads were visualized in IGV v2.17.4 with quick consensus mode and ‘hide small indels’ (below 50bp) options. Soft-clipped read segments were shown, unless mentioned otherwise.

For samples with read depth (DP) < 3 in the *SMN* PSV region in one or more haplotypes, haplotype phasing was improved by manual curation: first, ploidy-aware variant calling and phasing was performed on previously published SNV positions^20^ using a bed file containing these positions. Next, the phasing region was expanded by selection of SNVs upstream and downstream of known SNVs on positions where all haplotypes had minimum read depth of 4x and contained the variant in either 0% or 100% of the reads, allowing two errors in total and excluding variants located on homopolymer stretches of 5bp or more. The selected SNV positions were added to the bed file of known SNV positions and used for the next iterations. The resulting bed file was used for phasing in two SMA patients and six 1000G samples. An overview of phasing method per sample is included in **Table S1**. For each haplotype separately, SNV/INDEL variant calling was performed using Clair3 v1.0.4, and structural variant calling was performed using Sniffles2 v2.2.

### Variant filtering and visualization

Clair3 VCFs were parsed into tab-separated files that were merged into one file for all samples. Read depth was called for all samples on all variant positions with samtools v1.17, and added as an extra column to the variant files. The resulting file was loaded into R v4.4.0 for further analysis. Variants with read depth (DP) ≥3 and allele frequency (AF) ≥0.5 were categorized as alternative allele (ALT), DP≥3 and AF<0.5 (or not called by Clair3) as reference allele (REF), and DP<3 as not available (NA). All ALT positions were summarized and split per haplotype (**Table S2**). Next, FASTA files were made of each haplotype, using **Table S2** and a minimum read depth of 3x on every position on the phasing ROI called by samtools v1.17. The resulting FASTA files were concatenated per *SMN* copy type (*SMN1* or *SMN2*), which were mapped to the masked T2T-CHM13 reference genome using hapdiff v0.8 for visualization in IGV v2.17.4 without quick consensus mode and with the ‘hide small indels’ option disabled. Soft-clipped read segments were not shown. In 1000G samples, variants were classified as *SMN2*-specific when the alternative allele was present in 290% of resolved *SMN2* haplotypes and ≤20% of resolved *SMN1* haplotypes, based on at least 20 haplotypes each. Since PSV5 at position chr5:71,407,128 is a hybrid PSV in the T2T-CHM13 reference genome (cytosine instead of thymine, normally present in *SMN2*), it was classified as *SMN2*-specific when the alternative allele was present in ≤10% of resolved *SMN2* haplotypes and ≥80% of resolved *SMN1* haplotypes. At all *SMN2*-specific positions except PSV5, REF alleles were classified as *SMN1* PSV or environment SNV, and ALT alleles were classified as *SMN2* PSV or environment SNV (vice versa for PSV5). If an *SMN2* haplotype contained an *SMN1*-specific variant only at position chr5:71,407,825, previously reported as PSV8^15^, it was not considered a hybrid gene, because the genotypes at this position were highly variable in our patient population and was not consistently associated with other PSVs or *SMN1* environment markers in SMA haplotypes, in agreement with a recent study by Costa-Roger et al.^16^. To determine downstream *SMN1* environment SNV ratio, the number of haplotypes with downstream *SMN1* environment SNVs was divided by the total number of haplotypes in a sample; a mixed environment was counted as 0.5. If not all haplotypes were resolved in this region due to low read depth, the ratio was estimated with the allele frequency on the *SMN2*-specific positions in the full sample (ALT frequency of ∼0.67 results in *SMN1* environment ratio of 0.33). The presence of *NAIP* or truncated *NAIPP* gene in every haplotype was determined by visual inspection of read clipping as shown in **Fig. S2**, with a minimum requirement of three supporting reads.

### Comparison of ONT and Illumina variant calling

Illumina FASTQ data was trimmed for adapter sequences using trimmomatic v0.39 with the ILLUMINACLIP option and mapped to the masked T2T-CHM13 reference genome using BWAMEM2 v2.2.1. *SMN* ploidy-aware variant calling was performed by GATK v4.2.1.0 on the long-range PCR coordinates (T2T-CHM13 chr5:71,378,876-71,410,446). Variants were filtered for minimum depth (DP≥10) and minimum allele frequency (AF≥0.15, in which AF = (ALT-AD/DP)). The cutoff 0.15 was selected as this is just below the minimal expected allele frequency for the sample with the highest number of known *SMN* copies (5 copies, thus 0.20). Overlap between variants called by Illumina and ONT was considered per sample. A variant was considered present in both Illumina and ONT data when the variant was present in the Illumina GATK VCF (with DP≥10 and AF≥0.15) and at least one of the haplotype-specific ONT Clair3 VCFs (with DP≥3 and AF≥0.5).

### Annotation of possibly functional variants

For annotation of possibly functional variants, Clair3 VCFs of all SMA haplotypes were merged with vcftools v0.1.16 and converted to GRCh38 coordinates with the bcftools v1.18 liftover plugin. The Ensembl Variant Effect Predictor v111^44^ was used to assess the impact of genetic variants on canonical *SMN1* transcript ENST00000380707.9 with the following tools: SIFT, PolyPhen, AlphaMissense, CADD (PHRED≥15 considered possibly pathogenic) and SpliceAI (delta score≥0.5 considered possibly pathogenic). Only Clair3 variants with DP≥3 and AF≥0.5, that were also found with Illumina sequencing, were kept. Exonic and flanking intronic variants that were not predicted to have any effect, are also shown for completeness.

### Statistics

To test the difference between two groups of datapoints (**Fig. 1E-H**), an unpaired two-sided T-test was used. If the normality assumption, tested with the Shapiro-Wilk test, was violated, the Wilcoxon rank sum test was used. To test linear relationships, simple linear regression was performed. To test whether hybrid *SMN2* genes have a downstream *SMN1* environment more often than non-hybrid *SMN2* genes, a Fisher’s exact test was used. To test the difference in *SMN1* environment SNV ratio between disease severity groups, the Kruskal-Wallis rank sum test was used. Statistics were performed in R v4.4.0.

## Data availability

The raw sequencing data that support the findings of this study are available upon reasonable request from the corresponding authors (EJNG and GWvH). The adaptive sampling target file, variant calls at *SMN2*-specific positions and an overview of all Clair3 variant calls per haplotype are available in the supplementary materials.

## Code availability

The code for HapSMA is available at: https://github.com/UMCUGenetics/HapSMA. The code for analyses subsequent to HapSMA are available at: https://github.com/UMCUGenetics/ManuscriptSMNGeneConversion/tree/release_v1.0.0.

