## Supplementary information for "Long-read sequencing identifies copy-specific markers of *SMN* gene conversion in spinal muscular atrophy"

Contents

- Supplementary note 1
- Supplementary figures 1 – 7

Supplementary files

- Table_S1_SMN2_specific_variants_per_haplotype.xlsx (overview of *SMN2*-specific variants per haplotype)
- Table_S2_all_variants_per_haplotype.xslx (overview of all identified variants per haplotype, within the phasing ROI)
- adaptive_sampling_SMN_locus.fasta (target FASTA file for adaptive sampling)

**Supplementary note 1: sequence variants in the *SMN* gene**

Three missense, two synonymous and two untranslated region (UTR) variants were found in SMA patients (**Table 2**). Variant c.542A>G is described in the main text. Missense variant c.77G>A (p.Gly26Asp) is located in exon 1 of *SMN2*, near the Gemin2-binding domain of the SMN protein[1]. It is predicted to be pathogenic, and was found in *SMN2* in two patients with concordant and discordant (more severe) phenotype, making this variant a candidate negative modifier. A third missense variant, c.593C>T (p.Pro198Leu), was found in exon 4 in two out of four *SMN2* haplotypes of a discordant patient with SMA type 2b. Pathogenicity predictions from SIFT, PolyPhen and CADD and the relatively severe phenotype of this patient suggest that this variant might be a negative modifier. Synonymous variants c.84C>T and c.462A>G were not predicted to be pathogenic and were found in both concordant and discordant patients. 5' UTR variant c.-14C>T, intronic variant c.81+45C>T and 3' UTR variant c.*58G>A were found in the same haplotype of a concordant patient, and were not predicted to be pathogenic, whereas c.-14C>T and c.81+45C>T were previously reported in a different, severely affected, discordant patient[2]. Other intronic variants of unknown functional relevance, located in 100bp exon flanks are also reported (**Table 2**). No variants were found in the 100bp upstream of the translation start site, suggested to contain most critical promoter elements[3].

### Supplementary figures

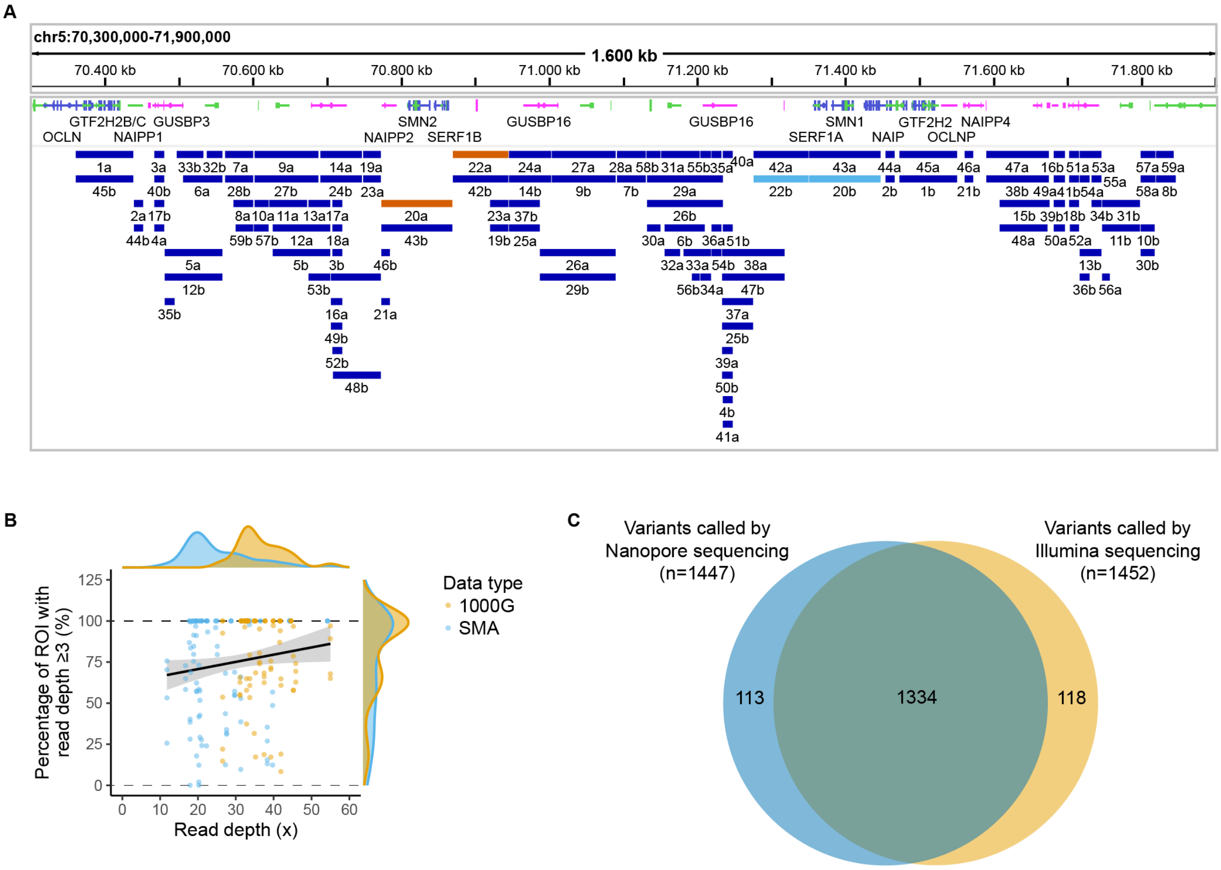


#### Supplementary figure 1: Performance of phasing and variant calling

**(A)** IGV snapshot of duplication segments on the *SMN* locus resulting from nucmer segmental duplication analysis of T2T-CHM13 chromosome 5 versus chromosome 5 (**Fig. 1A**). Duplicated segments share the same number. Segments 20a and 22a (chr5:70,772,138-70,944,284) were masked, whereas segments 20b and 22b (chr5:71,274,893-71,447,410) were used as ROI for phasing.

**(B)** Association between read depth and performance of haplotype phasing, with one datapoint per haplotype (n = 97 for 1000G and n = 104 for SMA). There is a small but significant association between total read depth on 30Mb region surrounding the *SMN* locus and the percentage of the phasing ROI with read depth ≥3 (simple linear regression, used model: percentage = 61.90 + 0.4398*(read depth), adjusted R-squared: 0.01643, F(1,199) = 4.34, p = 0.0385).

**(C)** Comparison of variant calling within the long-range PCR region by long-read sequencing (ONT) and short-read sequencing (Illumina). Out of all 1447 variants found by Nanopore sequencing across all samples sequenced with both methods (n = 29), 1334 were also found by Illumina sequencing (92.2%).

Out of all 1452 variants found by Illumina sequencing across all samples, 1334 were also found by Nanopore sequencing (91.9%).

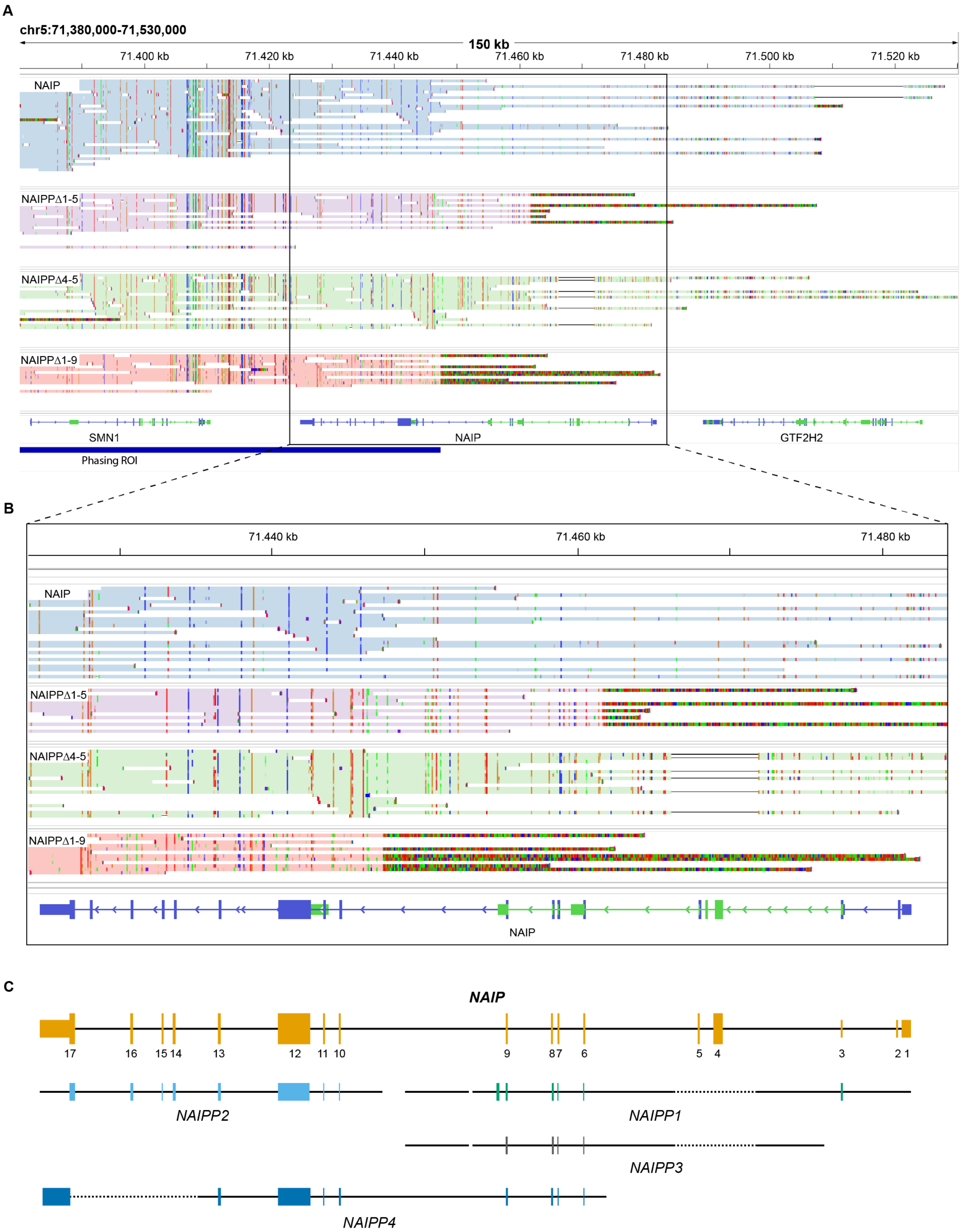


**Supplementary figure 2: The *SMN* genes can be followed by different *(pseudo)NAIP* genes**

**(A)** IGV snapshot of haplotypes in which different *NAIP* and *pseudoNAIP (NAIPP)* genes follow *SMN1/2*: full-length *NAIP*, with sequencing reads covering the entire gene; *NAIPPΔ4-5* missing exon 4-5;

*NAIPPΔ1-5* missing exon 1-5; and *NAIPPΔ1-9* missing exon 1-9. Deletions are shown by thin black lines, soft-clipping is shown as intensely, non-uniformely colored read segments.

**(B)** Zoomed in image of (A).

**(C)** Different *NAIPP* genes aligned to full-length *NAIP*, at scale with (B). *NAIP* exons are numbered, and protein-coding exons are displayed wider than untranslated regions. Black lines represent introns and flanks of the *NAIPP* genes, that align to the *NAIP* gene. Dashed lines represent parts that are missing in the *NAIPP* gene relative to the *NAIP* gene. *NAIPPΔ4-5* from (B) resembles the *NAIPP1* deletion; *NAIPPΔ1-5* resembles the *NAIPP4* breakpoint; and *NAIPPΔ1-9* resembles the *NAIPP2* breakpoint.

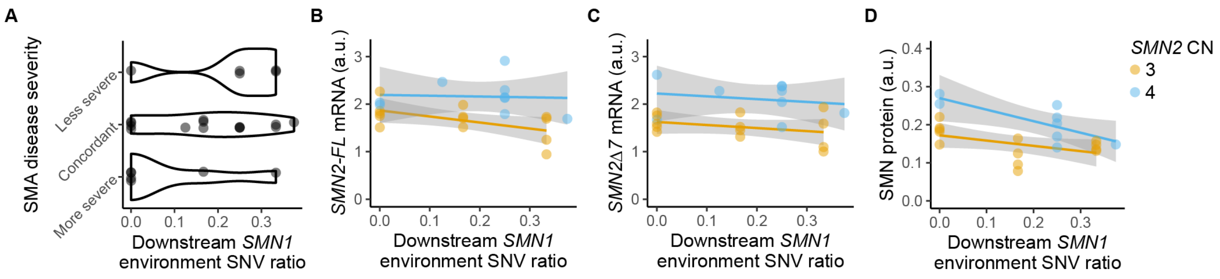


**Supplementary figure 3: Link between downstream *SMN1* environment SNV ratio and disease severity, mRNA expression and protein expression in patients with homozygous *SMN1* deletion and 3 or 4 *SMN2* copies**

Downstream *SMN1* environment SNV ratio was calculated as the number of haplotypes with downstream *SMN1* environment SNVs divided by total number of haplotypes.

**(A)** Comparison between disease severities. *SMN1* environment SNV ratio did not significantly differ between less severe (n = 5, median = 0.25), concordant (n = 14, median = 0.17), and more severe (n = 6, median = 0) patients (Kruskal-Wallis rank sum test, $\chi^2(2) = 3.141$, p = 0.208).

**(B)** *SMN2-FL* mRNA expression differed between copy groups but was not associated with downstream *SMN1* environment SNV ratio (simple linear regression, used model: expression = 0.2327 + 0.5254 * (copy group) - 0.8946 * (ratio), adjusted R-squared: 0.3748, F(2,19) = 7.294, p = 0.00446 for complete model; p = 0.00171 for copy group and p = 0.1111 for ratio).

**(C)** *SMN2Δ7* mRNA expression differed between copy groups but was not associated with downstream *SMN1* environment SNV ratio (simple linear regression, used model: expression = -0.1987 + 0.6069 * (copy group) - 0.6236 * (ratio), adjusted R-squared: 0.4578, F(2,19) = 9.866, p = 0.001152 for complete model; p = 0.000303 for copy group and p = 0.2380 for ratio).

**(D)** SMN protein expression differed between copy groups and was associated with downstream *SMN1* environment SNV ratio (simple linear regression, used model: expression = -0.02049 + 0.06704 * (copy group) - 0.19309 * (ratio), adjusted R-squared: 0.5081, F(2,18) = 11.33, p = 0.000653 for complete model; p = 0.000802 for copy group and p = 0.004746 for ratio).

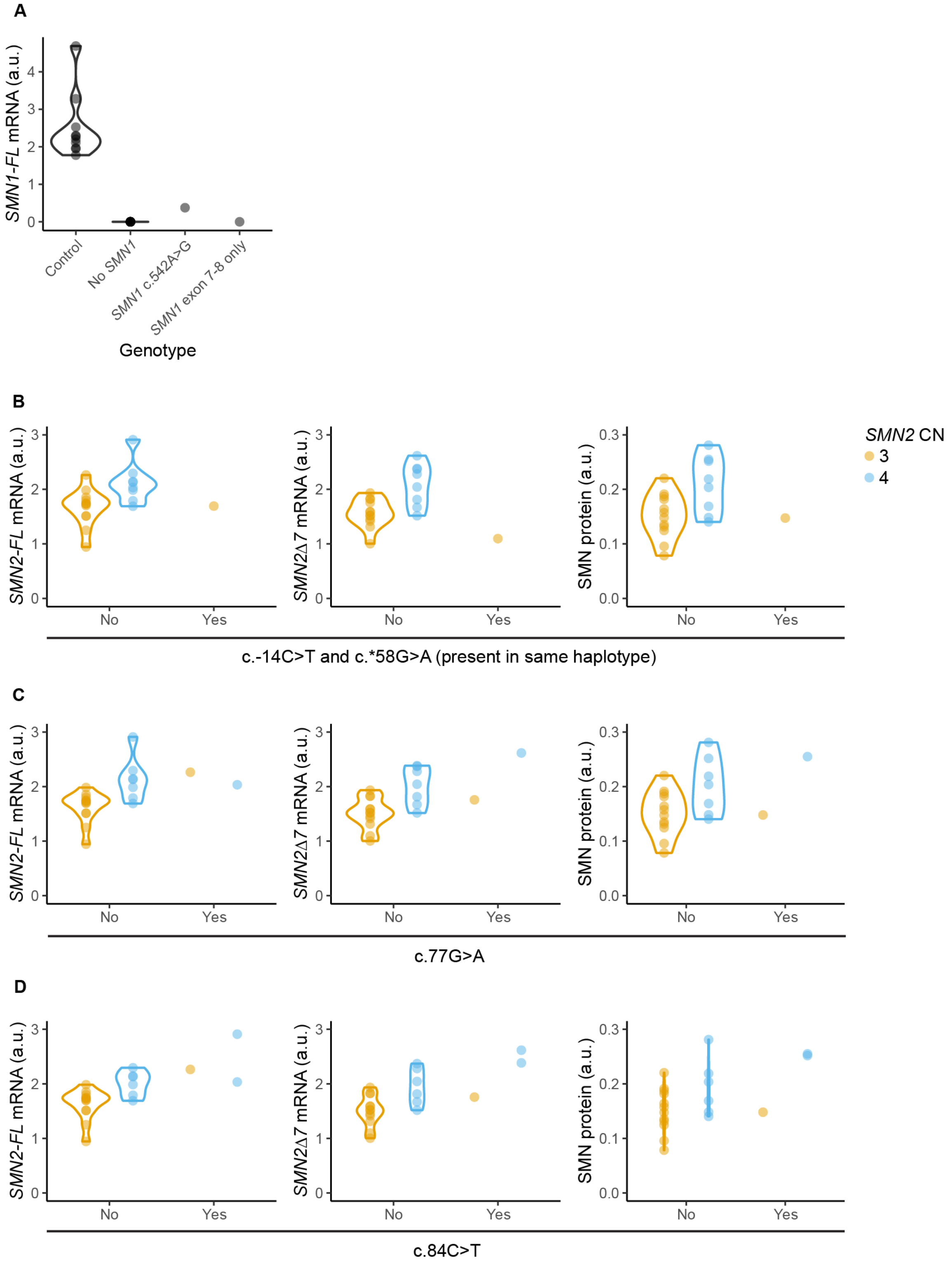


**Supplementary figure 4: Link between exonic variants and *SMN* mRNA and protein expression**

Each datapoint represents one individual. Only the variants for which expression data was available, are shown. Statistics were not performed due to small sample size per group. **(A)** *SMN1-FL* mRNA expression in fibroblasts of healthy controls, SMA patients with homozygous deletion of *SMN1*, one SMA patient with the c.542A>G variant in *SMN1* and one patient with only exon 7 and 8 of *SMN1*.

**(B-D)** mRNA and protein expression in fibroblasts of patients with exonic variants in *SMN2*. Variants c.-14C>T and c.*58G>A (B), c.77G>A (C) and c.84C>T (D) were present in only one haplotype per patient. CN: copy number; *SMN1-FL*: full-length *SMN1*; *SMN2-FL*: full-length *SMN2*; *SMN2Δ7*: *SMN2* lacking exon 7.

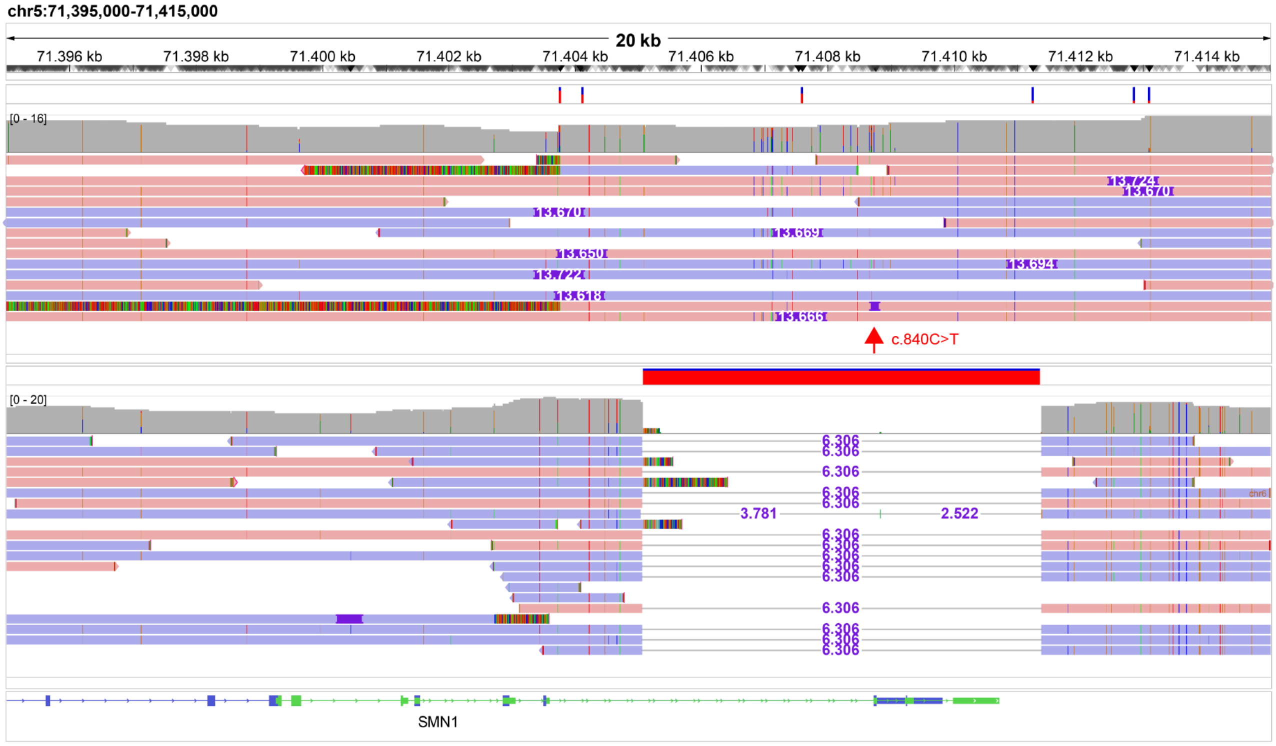


**Supplementary figure 3: Structural variants (SVs) in the *SMN* gene**

Each panel contains from top to bottom SV calls made with Sniffles2, read depth plot and sequencing reads colored by read strand. The upper panel shows sequencing reads of one haplotype from a sample with decreased copy number of exon 1-6 in MLPA. In most nanopore reads, this SV was detected as a ~13.7kb insertion containing exon 7 and 8 of *SMN1* or *SMN2*, depending on the location of the insertion upstream or downstream the c.840C>T PSV (marked with a red arrow). This indicates that *SMN1* exon 7 and 8 are inserted downstream of *SMN2*. However, it is possible that this structural variant is a deletion of exon 1-6 of *SMN1*, as found previously[4,5], extending to the downstream area of *SMN2,* if both genes were originally in the same orientation. Both situations result in an incomplete *SMN1* copy containing only exon 7 and 8. The insertion/deletion breakpoint, as can be seen from the soft-clipping in three reads, is located on Alu element AluYd8, which could be the mediator for this structural rearrangement. The lower panel shows a 6.3kb deletion of exon 7-8 of *SMN1/2*. Since this deletion covers the c.840C>T PSV, it was unknown whether this SV is present in an *SMN1* or *SMN2* copy; downstream *SMN2* environment SNVs suggest that it was originally an *SMN2* copy, as reported previously[6].

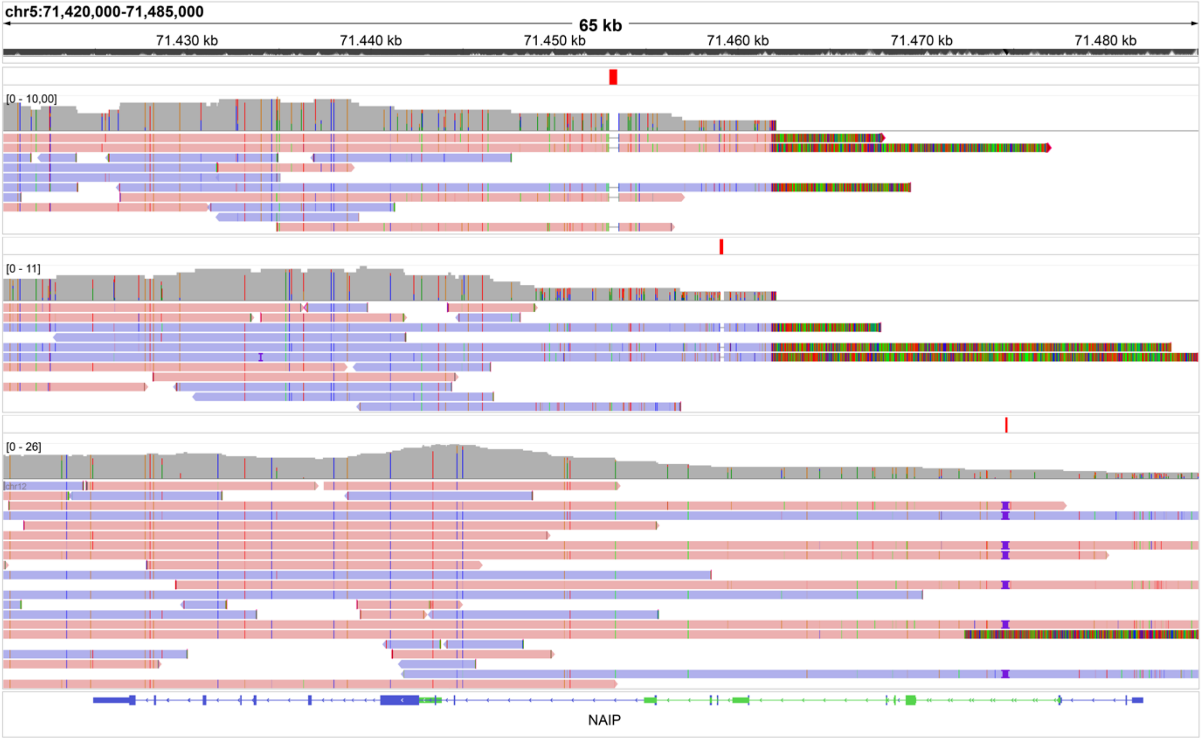


**Supplementary figure 4: SVs in the *(pseudo)NAIP* genes other than the truncations shown in supplementary figure 2**

Each panel contains from top to bottom SV calls made with Sniffles2, read depth plot and sequencing reads colored by read strand. Upper panel: a 519bp deletion was found in intron 9 of *NAIPΔ1-5* in 1 SMA haplotype. Middle panel: a 219bp deletion was found in intron 6 of *NAIPΔ1-5* in 2 SMA haplotypes. Lower panel: in two related 1000G samples, an AluYa5 insertion of ~330bp was found in *NAIP* intron 3, which was likely passed on from the mother to the child.

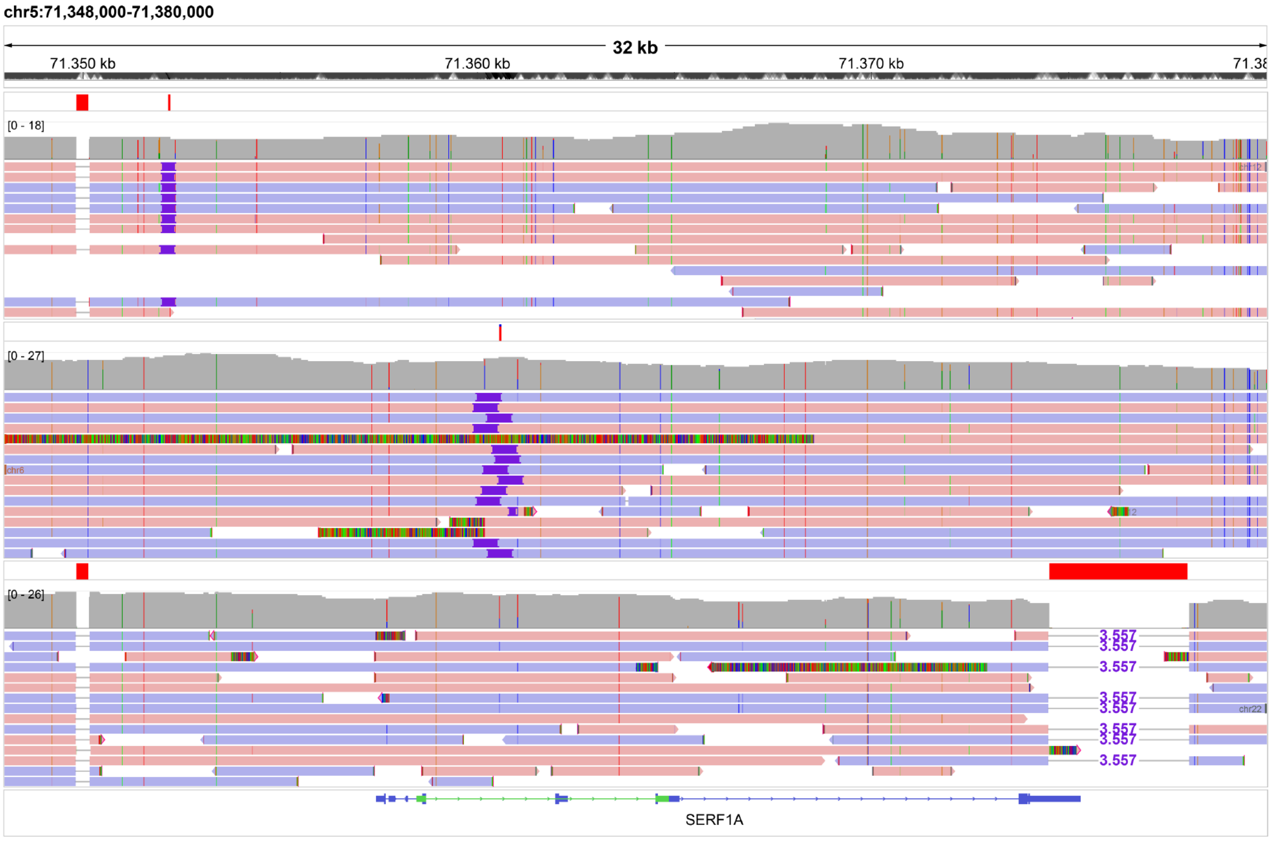


**Supplementary figure 5: SVs in and near the *SERF1A/B* gene**

Each panel contains from top to bottom SV calls made with Sniffles2, read depth plot and sequencing reads colored by read strand. Upper panel: Upstream of *SERF1A/B*, a 346bp AluYb8 deletion was found in 38 SMA and 60 1000G haplotypes, and a ~330bp AluYa5 insertion in 13 SMA and six 1000G haplotypes. The insertion was only found in haplotypes where the deletion was also present. Middle panel: in intron 2 of *SERF1A/B*, one 1000G haplotype had a 612bp insertion. Lower panel: four 1000G haplotypes had a 3557bp deletion near the last exon of the long transcript of *SERF1A/B*.
